# COV-ID: A LAMP sequencing approach for high-throughput co-detection of SARS-CoV-2 and influenza virus in human saliva

**DOI:** 10.1101/2021.04.23.21255523

**Authors:** Robert Warneford-Thomson, Parisha P. Shah, Patrick Lundgren, Jonathan Lerner, Benjamin S. Abella, Kenneth S. Zaret, Jonathan Schug, Rajan Jain, Christoph A. Thaiss, Roberto Bonasio

## Abstract

The COVID-19 pandemic has created an urgent need for rapid, effective, and low-cost SARS-CoV-2 diagnostic testing. Here, we describe COV-ID, an approach that combines RT-LAMP with deep sequencing to detect SARS-CoV-2 in unprocessed human saliva with high sensitivity (5–10 virions). Based on a multi-dimensional barcoding strategy, COV-ID can be used to test thousands of samples overnight in a single sequencing run with limited labor and laboratory equipment. The sequencing-based readout allows COV-ID to detect multiple amplicons simultaneously, including key controls such as host transcripts and artificial spike-ins, as well as multiple pathogens. Here we demonstrate this flexibility by simultaneous detection of 4 amplicons in contrived saliva samples: SARS-CoV-2, influenza A, human *STATHERIN*, and an artificial SARS spike-in. The approach was validated on clinical saliva samples, where it showed 100% agreement with RT-qPCR. COV-ID can also be performed directly on saliva adsorbed on filter paper, simplifying collection logistics and sample handling.

## INTRODUCTION

Within the first year of the COVID-19 pandemic SARS-CoV-2 has swept across the world, leading to more than 130 million infections and over 2.8 million deaths worldwide (as of April 2021). In many countries, non-pharmaceutical interventions, such as school closures and national lockdowns, have proven to be effective, but could not be sustained due to economic and social impact^1,2^. Regularly performed population-level diagnostic testing is an attractive solution^3^, particularly as asymptomatic individuals are implicated in rapid disease transmission, with a strong overdispersion in secondary transmission^4^. Maintenance of population-level testing can be successful in isolating asymptomatic individuals and preventing sustained transmission^5, 6^; however, considerable barriers exist to the adoption of such massive testing strategies. Two such barriers are cost and supply constraints for commercial testing reagents, both of which make it impractical to test large numbers of asymptomatic individuals on a recurrent basis. A third major barrier is the lack of “user-friendly” protocols that can be rapidly adopted by public and private organizations to establish high-throughput surveillance screening. In addition, while COVID-19 testing of symptomatic individuals might be effective during the summer season, when other respiratory infections are rare, new strategies are needed to facilitate rapid differential diagnosis between SARS-CoV-2 and other respiratory viruses in winter.

Recent adaptations of reverse transcription and polymerase chain reaction (RT-PCR) to amplify viral sequence and perform next-generation DNA sequencing have opened promising new avenues for massively parallel SARS-CoV-2 detection. In general, sequencing-based protocols use libraries of amplification primers to tag reads originating from each individual patient sample with a unique index that can be identified and deconvoluted after sequencing, thus allowing pooling of tens of thousands of samples in a single assay. SARSeq, SPAR-Seq, Swab-seq, and INSIGHT, directly amplify the viral RNA by RT-PCR and simultaneously introduce barcodes^7-10^. While effective, these methods rely on individual PCR amplification of each patient sample, thus requiring a large number of thermal cyclers for massive scale-up. An alternative approach, ApharSeq, addresses this bottleneck by annealing barcoded RT primers to viral RNA and pooling samples prior to amplification but the need for specialized oligo-dT magnetic beads might constitute a separate adoption barrier for this method^11^. Finally, several recent methods have been designed to take advantage of the extreme sensitivity and isothermal conditions of loop-mediated isothermal amplification (LAMP)^12-14^, but these methods either require additional manipulation to introduce barcodes^12, 13^ or do not allow for convenient multiplexing^14^.

In this study, we present COV-ID, a method for SARS-CoV-2 identification based on LAMP, which enables large-scale diagnostic testing at low cost and with minimal on-site equipment. COV-ID is a robust method that can be used to test tens of thousands of samples for multiple pathogens with modest reagent costs and 2–4 laboratory personnel, generating results within 24 hours. COV-ID uses unpurified saliva or saliva adsorbed on filter paper as input material, thus enabling the massively parallel, inexpensive testing required for population-level surveillance of the COVID-19 pandemic (**Fig. 1A**).

**Figure 1.**
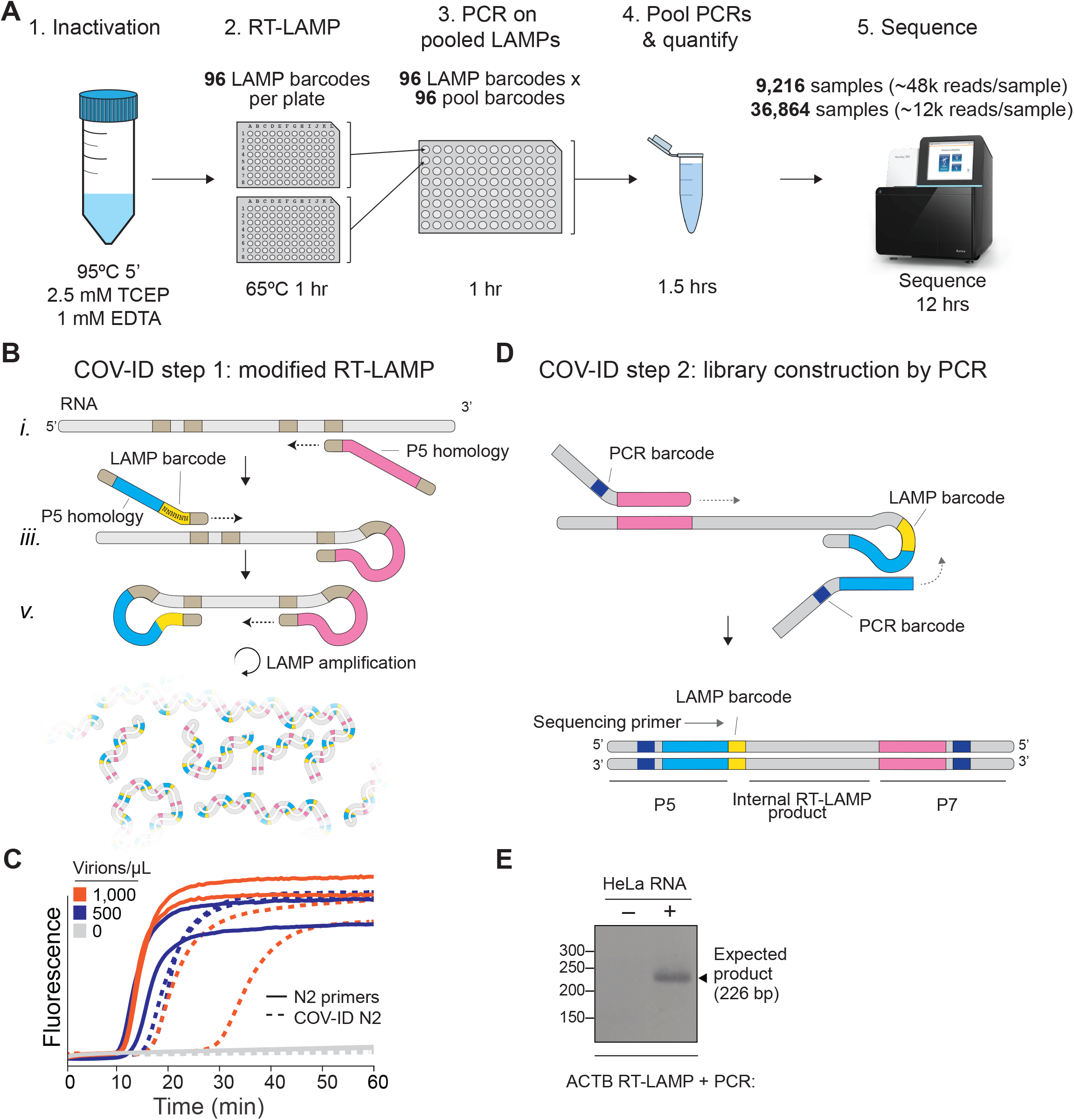
Barcoding and PCR amplification of RT-LAMP products. (A) Overview of COV-ID. Saliva is collected and inactivated prior to RT-LAMP performed with up to 96 individual sample barcoded primers. LAMP reactions are pooled and further amplified via PCR to introduce Illumina adapter sequences and pool-level dual indexes. A single thermal cycler can amplify 96 or 384 such pools and the resulting “super-pool” can be sequenced overnight to detect multiple amplicons from 9,216 or 36,864 individual patient samples (number of reads in parenthesis assume an output of ∼450M reads from a NextSeq 500). (B) Schematic of the RT-LAMP (step I) of COV-ID. Selected numbered intermediates of RT-LAMP reaction are shown to illustrate how the LAMP barcode, shown in yellow, and the P5 and P7 homology sequences (blue and pink, respectively) are introduced in the final LAMP product. Upon generating the dumb-bell intermediate the reaction proceeds through rapid primed and self-primed extensions to form mixture of various DNA amplicons containing sequences for PCR amplification. A more detailed version of the LAMP phase of COV-ID, including specific sequences, is illustrated in Fig. S1. (C) Conventional RT-LAMP primers (solid lines) or primers modified for COV-ID (dotted lines) were used for RT-LAMP of SARS-CoV-2. The numbers of inactivated SARS-CoV-2 virions per µL is indicated in the color legend. (D) Schematic of the PCR (step II) of COV-ID. Following RT-LAMP, up to 96 reactions are pooled and purified and Illumina libraries are generated directly by PCR with dual-indexed P5 and P7 adapters in preparation for sequencing. (E) COV-ID primers targeting ACTB mRNA were used for RT-LAMP with HeLa total RNA. LAMP was diluted 1:100, amplified via PCR and resolved on 2% agarose gel.

## RESULTS

### Two-step amplification and indexing of viral and human sequences via RT-LAMP and PCR

The molecular basis for COV-ID is reverse transcription loop-mediated isothermal amplification (RT-LAMP), an alternative to PCR that has been used extensively for viral DNA or RNA detection in clinical samples^15-18^, including SARS-CoV-2^19, 20^. RT-LAMP requires 4–6 primers that recognize different regions of the target sequence^21, 22^ and proceeds through a set of primed and self-primed steps to yield many inverted copies of the target sequence spanning a range of molecular sizes (**Fig. S1**). The forward inner primer (FIP) and backward inner primer (BIP), which recognize internal sequences, are incorporated in opposite orientation across the target sequence in the final amplified product (**Fig. S1**).

Previous studies have shown that the FIP and BIP tolerate insertion of exogenous sequence between their different target homology regions^23^. We exploited this flexibility and introduced 1) patient-specific barcodes as shown previously^12, 14, 23^ and 2) artificial sequences that allowed for PCR amplification of a small product compatible with Illumina sequencing library construction (**Fig. 1, Fig. S1**). These innovations allow us to pool individually barcoded RT-LAMP reactions and amplify them in batch via PCR, while introducing unique P5 and P7 dual indexes in different pools, thus enabling two-dimensional barcoding and dramatically increasing method throughput (see **Table S1** for PCR primer sequences). To minimize pool variability, PCR primers can be titrated to 100 nM and pooled PCRs carried out to completion, resulting in each pool being amplified to the same approximate concentration. Uniquely amplified and barcoded pools are mixed into a single “super-pool” that can be sequenced on an Illumina NextSeq or similar instrument (**Fig. 1A**). Combining individual barcodes embedded in the product at the RT-LAMP step with dual indexes introduced at the pool level during the PCR step allows for deconvolution of thousands or tens of thousands of samples in a single sequencing run.

To determine whether introduction of these exogenous sequences into the primers inhibited the isothermal amplification step, we performed RT-LAMP on inactivated SARS-CoV-2 virus using an extensively validated primer set against the N2 region of the nucleocapsid protein^24^ including either the conventional BIP and FIP primers or their modified version re-engineered for the COV-ID workflow (**Fig. 1B**). Although the appearance of the amplified viral product was slightly delayed when using COV-ID primers, all reactions reached saturation rapidly and without detectable amplification of negative controls (**Fig. 1C**). Next, we tested whether COV-ID was compatible with RT-LAMP using newly designed primers against a host (human) transcript and whether the second step of COV-ID, direct library construction and indexing via PCR amplification (**Fig. 1D**), yields the desired product. For this, we designed RT-LAMP primers against the human beta-actin (*ACTB*) transcript that included sequences necessary for COV-ID. After RT-LAMP, reactions were diluted 100-fold before PCR with barcoded Illumina adapters. A PCR product of the expected size was visible in reactions containing total HeLa RNA, whereas no PCR product was observed in the absence of template (**Fig. 1E**). Sanger sequencing of the PCR product confirmed that RT-LAMP followed by PCR generated the product expected by the COV-ID method design, including the sample barcode introduced during the RT-LAMP step. Thus, our data show that RT-LAMP is tolerant of sequence insertions in the BIP and FIP primers that allow introduction of LAMP-level barcodes as well as sequences homologous to Illumina adapters for direct amplification, indexing, and library construction via PCR.

### Sequencing-based detection of SARS-CoV-2 RNA from saliva using COV-ID

We next evaluated the utility of COV-ID to detect viral RNA in saliva. Saliva is an attractive sample material for COVID-19 diagnostics with potential for early detection^25^, and has been shown to be a viable template for nucleic acid amplification via RT-PCR^26^, recombinase polymerase amplification (RPA)^27^ as well as RT-LAMP^28, 29^. We prepared human saliva for RT-LAMP using a previously described treatment that inactivates SARS-CoV-2 virions, saliva-borne RNases and LAMP inhibitors (**Fig. 2A**)^29^. We performed RT-LAMP followed by PCR on inactivated saliva spiked with water or 1,000 total copies of inactivated SARS-CoV-2 virus. We observed a single band of the expected size in reactions performed on saliva spiked with virus but not in control reactions (**Fig. 2B**). The sequence of the amplified and barcoded viral product was confirmed by Sanger sequencing (**Fig. S2A**). Next, we subjected the libraries to deep sequencing. Reads aligned uniformly to the *N* gene, the region targeted by the N2 primer set, in COV-ID libraries constructed from viral samples but not in control libraries (**Fig. 2C**).

**Figure 2.**
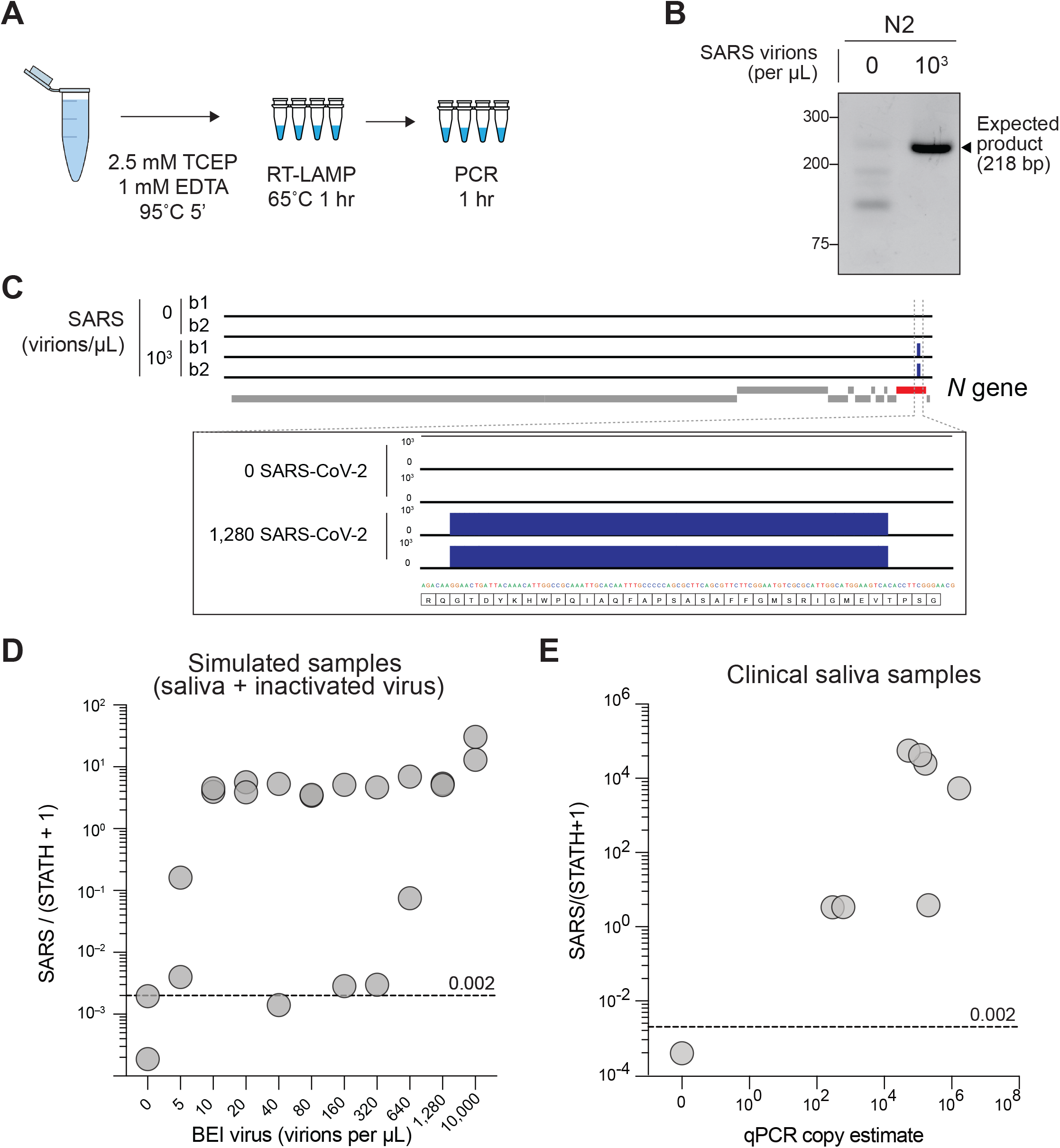
Sequencing-based detection of SARS-CoV-2 in saliva samples. (A) Saliva preparation. Crude saliva was inactivated via TCEP/EDTA addition and 95ºC incubation prior to RT-LAMP. (B) RT-LAMP followed by COV-ID PCR performed directly on saliva. Saliva with and without addition of 1,000 copies of inactivated SARS-COV-2 templates was inactivated as described in (A), then used as template. (C) Alignment of sequenced reads against SARS-COV-2 genome from COV-ID of inactivated saliva spiked with without 1,280 virions SARS-COV-2 per µL. All SARS-COV-2 reads align exclusively to expected region of the N gene. Open reading frames of viral genome are depicted via gray boxes below alignment. Inset: scale shows reads per 1,000. (D) Scatter plot for the ratio of SARS-CoV-2 / (*STATH* + 1) reads obtained by COV-ID (*y* axis) versus the number of virions per µL spiked in human saliva (x axis). The threshold was set above the highest values scored in a negative control (dashed line). (E) COV-ID performed on clinical saliva samples. The scatter plot shows the SARS-CoV-2 / (*STATH* + 1) read ratio (*y* axis) versus the viral load in the sample estimated by a clinically approved, qPCR-based diagnostic test. The threshold was set based on the negative controls shown in (D).

In several SARS-CoV-2 FDA approved tests, parallel amplification of a host (human) amplicon is implemented as a metric for sample integrity and quality. That is, if no human RNA is amplified from a clinical sample, no conclusion can be drawn from a negative test result^30^. However, in most tests, viral and human amplicons must be detected separately, resulting in a multiplication of the number of reactions to be performed. We reasoned that the deep sequencing nature of COV-ID would allow for simultaneous detection of viral, human, and other control amplicons, without increasing the number of necessary reactions. In fact, given that the PCR handles inserted in the BIP and FIP are the same for all RT-LAMP amplicons (**Fig. 1B**), the same P5 and P7 Illumina primers allow the simultaneous amplification of all RT-LAMP products obtained with COV-ID-modified primer sets (**Fig. 1D**). To identify a suitable human control, we compared conventional RT-LAMP primers for the mRNA of *ACTB*^24^ or *STATHERIN* (*STATH*), a gene expressed specifically in saliva^31^. To determine which of the two RT-LAMP primer sets was a better proxy to measure RNA integrity in saliva samples, we assayed for amplification of the respective products in presence or absence of RNase. Whereas addition of RNase A abolished the *STATH* signal, it was ineffectual for *ACTB* (**Fig. S2B**), suggesting that amplification of genomic DNA made considerable contributions to the RT-LAMP signal observed for the latter. Therefore, we utilized *STATH* mRNA as a human control in subsequent experiments.

We used COV-ID-adapted primer sets for *N2* and *STATH* (**Table S1**) in multiplex on inactivated saliva spiked with a range of SARS-CoV-2 from 5 to 10,000 virions/μL. Subsequently, each RT-LAMP reaction was separately amplified via PCR using a unique P5 and P7 index combination, pooled, quantified, and deep-sequenced to an average depth of 6,000 reads per sample. After read trimming, alignment, and filtering (see Methods), 76% of reads from saliva COV-ID reactions were informative (**Fig. S2C**). In order to differentiate SARS-CoV-2 positive and negative samples, we calculated the ratio between N2 reads and reads mapping to the human *STATH* control. Using the highest *N2*/*STATH* read ratio in control (SARS-CoV-2 negative saliva) as a threshold, 95% (19/20) of samples with spiked-in virus were correctly classified as positives (**Fig. 2D**). Using COV-ID, we consistently detected SARS-CoV-2 in saliva samples containing as low as 5 virions per µL, a sensitivity comparable and in some cases superior to those of established testing protocols^32^.

Scaling COV-ID to handle higher sample numbers requires pooling samples immediately following RT-LAMP, prior to the PCR step (**Fig. 1A**). We designed 32 unique 5-nucleotide barcodes for several target LAMP amplicons (**Fig. S2D** and **Table S2**). We first individually validated each barcode and primer combination by real-time fluorescence and PCR efficiency. Certain barcodes inhibited the RT-LAMP reaction, possibly due to internal micro-homology and primer self-hybridization^33^. Nonetheless, out of 32 barcodes tested in 3 separate RT-LAMP reactions (*N2, ACTB*, and *STATH*), 25 successfully amplified all three target RNAs (**Fig. S2D**). Saliva samples spiked with various concentrations of inactivated SARS-CoV-2 were amplified via barcoded RT-LAMP, then optionally pooled prior to PCR and sequencing (**Fig. S2E**). CoV-2/STATH ratios demonstrated no loss of sensitivity or specificity in the pooled samples compared to the individual PCRs.

To test the potential of COV-ID on patient samples, we tested saliva specimens, collected and previously analyzed at the Hospital of the University of Pennsylvania (see Methods). We carried out multiplex barcoded RT-LAMPs on each sample (COV-ID step I, **Fig. 1B**), pooled the reactions and then constructed libraries via PCR (COV-ID step II, **Fig. 1D**). After deep sequencing, analysis of *N2/STATH* ratios showed 100% (8/8) concordance with viral copy numbers generated by a standard clinical test (RNA purification followed by RT-qPCR) (**Fig. 2E**), demonstrating the effectiveness of the COV-ID approach.

Taken together, our data show that COV-ID can be utilized to detect viral and human amplicons in multiplex directly from saliva. The samples that can be batch amplified and deconvoluted after deep sequencing.

### Calibration of COV-ID using an artificial spike-in

Existing deep sequencing approaches for massively parallel COVID-19 testing based on RT-PCR incorporate artificial spike-ins, which serve as an internal calibration controls and allow for better estimates of viral loads by end-point PCR^7, 8^. At the same time, adding to the reactions an artificial substrate for amplification helps minimizing spurious signals as it can “scavenge” viral amplification primers in negative samples. Finally, by providing a baseline amplification even in empty samples, a properly designed spike-in strategy can reduce variance in total amounts of final amplified products across samples, which compresses the dynamic-range of sequence coverage for each patient in a complex pool and, therefore, reduces the risk of inconclusive samples due to low sequencing coverage^8^.

We reasoned that a spike-in approach for LAMP-based quantification would provide similar benefits in the context of COV-ID. To generate a SARS-CoV-2 spike-in, we synthesized a fragment of the *N2* RNA that retained all primer-binding regions for RT-LAMP and contained a divergent 7-nt stretch of sequence to distinguish reads originating from the spike-in from those originating from the natural virus (**Fig. S3A**). After confirming that the spike-in template was efficiently amplified via RT-LAMP with the *N2* primer set (**Fig. S3B**), we performed pooled COV-ID on virus-containing saliva in the presence of 20 fg of *N2* spike-in RNA. As expected^8^, addition of a constant amount of viral spike-in across reactions reduced the variability in total read numbers for individual samples in the final pool (**Fig. S3C**). As discussed above, a narrower range in sequencing output across samples in a pool optimizes the utilization of sequencing reads, and ultimately lowers the cost per sample. Because the spike-in provides an internal calibration that is independent of the RNA quality found in saliva, in several cases normalization against the spike-in resulted in lower levels of false positive signal from negative samples (**Fig. S3D**). This is likely because in cases where very few *STATH* reads were obtained, possibly due to degradation of host RNA in the saliva sample, the resulting small denominator inflated the *N2*/*STATH* ratio even for SARS-CoV-2 signal that was low in absolute terms and likely spurious.

Thus, these data show that spike-in strategies are compatible with the COV-ID workflow and provide a means to stabilize total amplification and read allocation per sample while also offering an additional calibration control to better estimate the viral load in samples where the endogenous *STATH* mRNA might be below detection due to improper collection or handling.

### Simultaneous detection of SARS-CoV-2 and influenza A by COV-ID

Given the challenge of distinguishing early symptoms of COVID-19 from other respiratory infections, we evaluated COV-ID for the simultaneous detection of more than one viral pathogen. Multiple distinct products can be simultaneously amplified by RT-LAMP in the same tube by providing the appropriate primer sets in multiplex, as we demonstrated above by co-amplifying *N2* and *STATH* in the same COV-ID reaction (see **Fig. 2**). In fact, simultaneous detection of SARS-CoV-2 and influenza virus by RT-LAMP was previously achieved, albeit in a fluorescent-based, low-throughput type of assay^34^. We reasoned that the sequencing-based readout of COV-ID would allow extending this approach to the simultaneous detection of multiple pathogens as well as endogenous (host mRNA) and artificial (spike-in) calibration standards, all in a single reaction.

To test the ability of COV-ID to simultaneously detect multiple viral templates, we selected and validated a generic “flu” RT-LAMP primer set that recognizes several strains, including influenza A virus (IAV) and influenza B^34, 35^, and modified the BIP and FIP sequence to introduce the COV-ID barcodes and handles for PCR (**Fig. S2D** and **Table S1**). We added inactivated SARS-CoV-2 virus (BEI resources) and IAV strain H1N1 RNA (Twist Biosciences) to saliva according to a 3 × 4 matrix of (10^4^, 10^3^, or 0 copies per µL) SARS-CoV-2 RNA against H1N1 RNA (10^5^, 10^4^, 10^3^, or 0 copies per µL) (**Fig. 3A**), as well as the *N2* spike-in control. We performed multiplex COV-ID on these samples using primers sets for *STATH, N2* (to detect SARS-CoV-2), and IAV (to detect H1N1) and sequenced to an average depth of 21,000 reads per sample. Both H1N1 and SARS-CoV-2 were detected above background and the signal correlated with the amount of the respective template added to saliva (**Fig. 3B–C**). Overall, multiplex COV-ID correctly identified samples that contained only SARS-CoV-2 (7/8) or H1N1 (6/8). For samples that contained both pathogens we observed reduced sensitivity (11/16 identification of both pathogens), which was also observed in a previous multiplexing attempt^34^. However, in practice individuals who are simultaneously infected with both viruses presumably would be rare, and for these cases the ability to detect at least one virus successfully would allow to follow up with further diagnostic testing. We found that of the samples containing both viruses, 16/16 showed positive detection of at least one pathogen (SARS-CoV-2 or H1N1), suggesting the reduced sensitivity of the multiplex assay is due to interference between amplification of both viral templates. This also demonstrates that COV-ID can be used as an effective screening approach for multiple viral templates.

**Figure 3.**
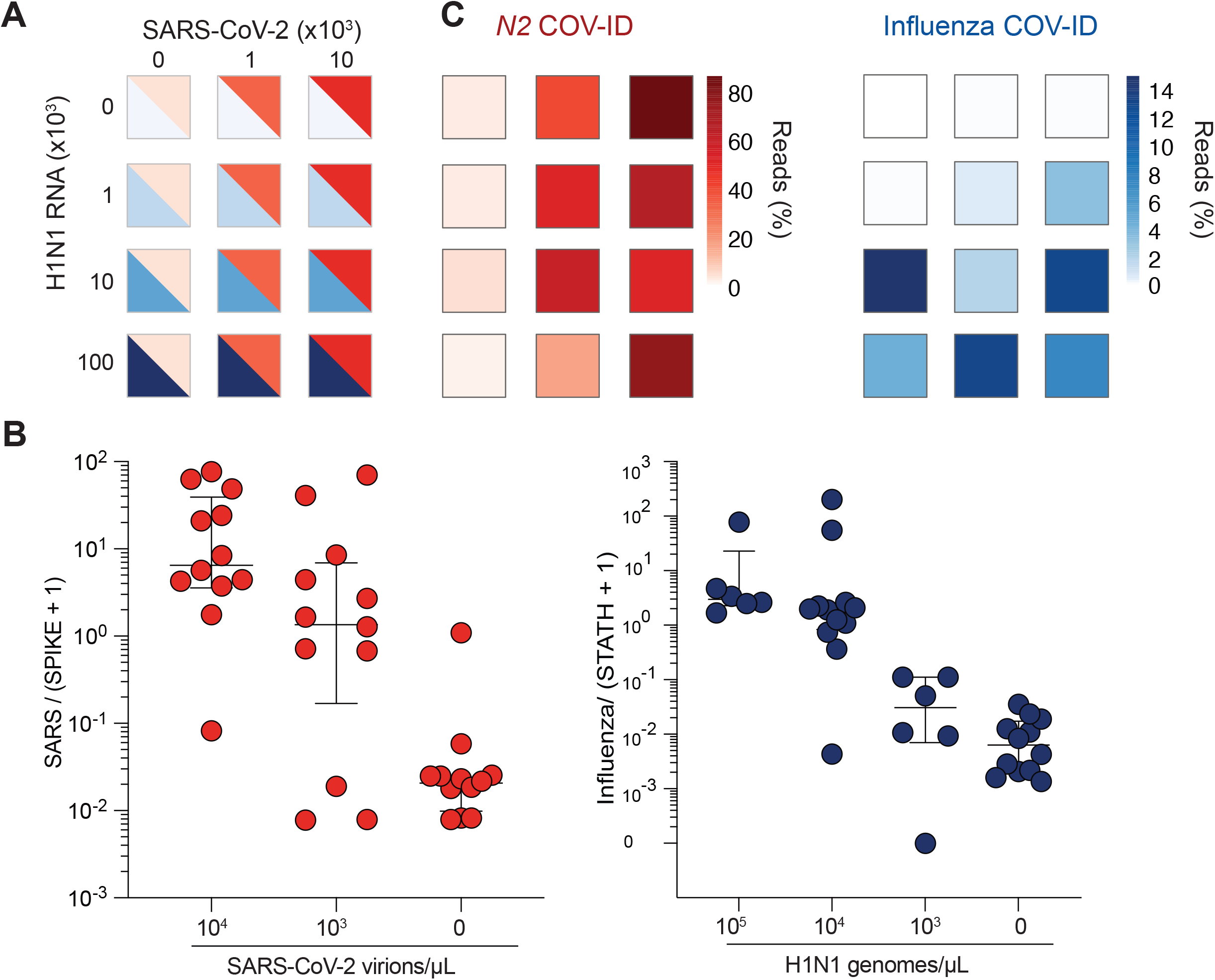
COV-ID multiplex detection of SARS-COV-2 and Influenza A. (A) TCEP/EDTA treated saliva was spiked with indicated amounts of BEI heat-inactivated SARS-CoV-2 or H1N1 influenza A RNA to the indicated concentration of virions/genomes per µL. 1 µl of saliva was used for COV-ID reactions. (B) COV-ID was performed in two independent experiments on saliva samples from the matrix shown in (A) in the presence of 20 femtograms synthetic N2 spike-in using N2, influenza (Zhang and Tanner, 2020) and STATH COV-ID primers. N2/(N2 Spike + 1) and influenza/(STATH + 1) read ratios are displayed with bars showing median ± interquartile range. (C) Heatmaps of SARS-CoV-2 (left) or H1N1 (right) COV-ID signal in multiplex reaction. Heatmaps are colored according by percentage of viral reads observed.

### Paper-based saliva sampling for COV-ID

As an additional step toward increasing the throughput of the COV-ID approach, we explored avenues to simplify collection, lower costs, and expedite processing time. Absorbent paper is an attractive alternative to sample vials for collection, given its low cost, wide availability, and smaller environmental footprint. In fact, paper has been used as a means to isolate nucleic acid from biological samples for direct RT-PCR testing^36^ as well as RT-LAMP^37, 38^.

We sought to determine whether the COV-ID workflow would be compatible with saliva collection on absorbent paper. First, we immersed a small square of Whatman filter paper into water containing various dilutions of inactivated SARS-CoV-2. After 2 min, the paper was removed and transferred to PCR strip tubes followed by heating at 95ºC for 5 minutes to air-dry the sample (**Fig. 4A**). Next, we added the RT-LAMP mix containing the *N2* COV-ID primer set directly to the tubes containing the paper squares and let the reaction proceed in the usual conditions. COV-ID PCR products of the correct size were evident in all samples containing viral RNA, with sensitivity of at least 100 virions / μL (**Fig. 4B**) and in none of the controls, demonstrating that the presence of paper does not interfere with the RT-LAMP reaction and subsequent PCR amplification with Illumina adapters.

**Figure 4.**
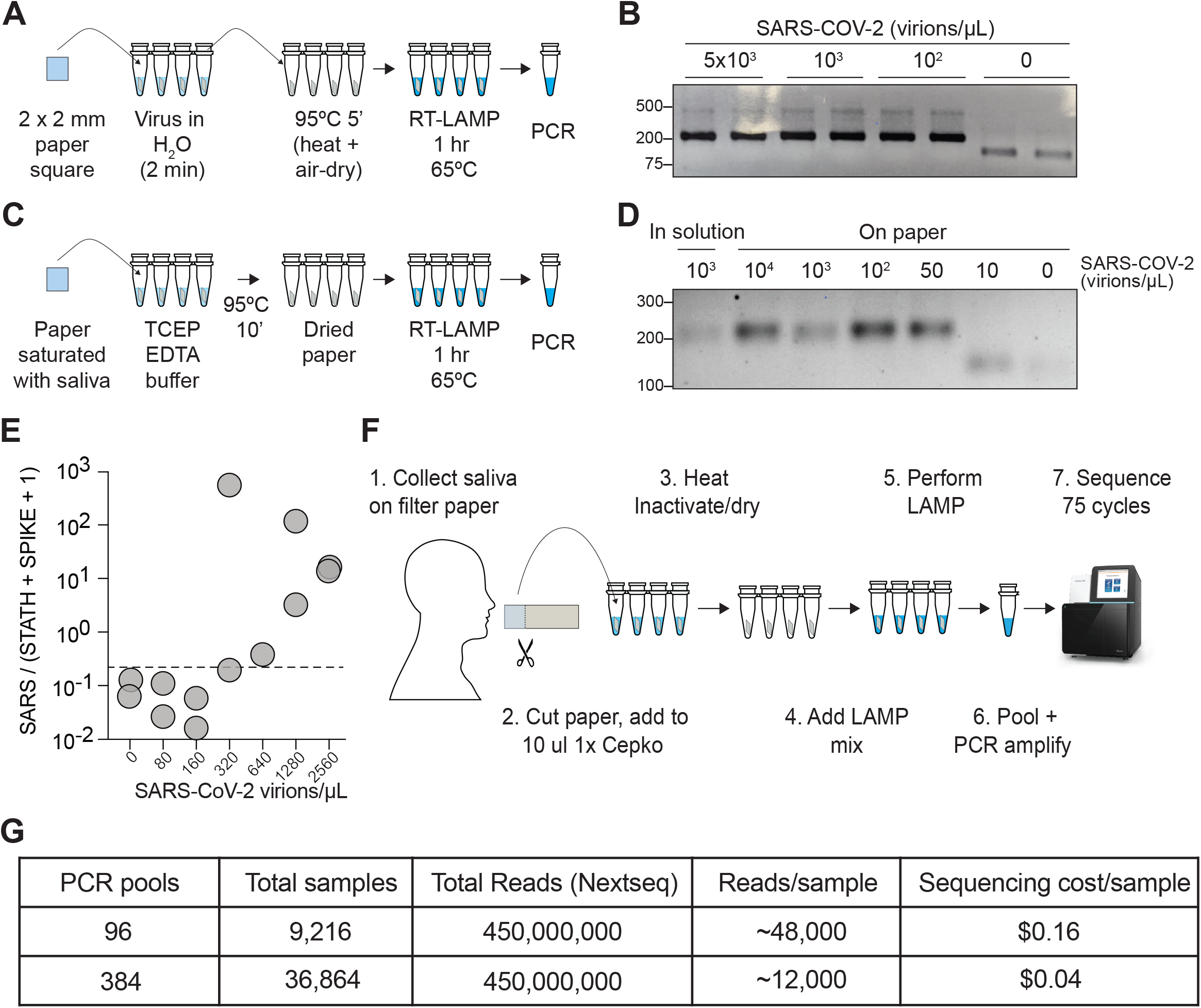
COV-ID on saliva collected on paper. (A) Scheme for COV-ID on viral RNA absorbed on paper. (B) PCR reactions from paper samples immersed in water with indicated viral concentrations then amplified with N2 COV-ID primers. (C) Scheme for COV-ID on saliva spiked with viral and RNA and absorbed on paper. (D) Same as (B) but on saliva absorbed on paper. (E) SARS-CoV-2 virus was added to saliva and prepared as in (C). RT-LAMP and sequencing was carried out in presence of SARS spike-in RNA. Viral reads are presented as ratio against the sum of STATH and N2 spike-in reads. Positive threshold was set as 2x maximum value in negative saliva and indicated by dashed horizontal line. (F–G): Paper-based COV-ID workflow (F) and cost calculations (G). Saliva is collected orally on a precut strip of paper, from which a 2 mm square would be cut out and added to a reaction vessel containing TCEP/EDTA inactivation buffer and processed as shown in (C).

To assay direct COV-ID detection from saliva on paper, we saturated Whatman filter paper squares with saliva containing different amounts of inactivated SARS-CoV-2 virus, which, we reasoned, would be equivalent to a patient collecting their own saliva by chewing on a small piece of absorbent paper. Next, we placed the paper squares into reaction tubes containing TCEP/EDTA inactivation buffer (see Methods) similar to that used for the in-solution samples used in our previous experiments (see **Fig. 1A**). We dried the paper at 95ºC and performed RT-LAMP followed by PCR (**Fig. 4C**), which resulted in the appearance of COV-ID products of the correct size starting from saliva spiked with as few as 50 virions / μL (**Fig. 4D**). We then performed COV-ID sequencing on saliva collected on paper using primers N2 and *STATH* in the presence of the *N2* spike-in RNA. The sequence data showed more variability and limited coverage of the control amplicons compared to in-solution COV-ID likely due to the more challenging reaction conditions; therefore, we normalized viral reads using both *STATH* and spike-in. This paper-based COV-ID proof-of-principle experiment detected the presence of viral RNA in samples with as little as 320 copies / µL (**Fig. 4E**), a lower sensitivity compared to that of in-solution COV-ID but still well within the useful range^39^ to detect infections.

Taken together, these data show that the RT-LAMP step of COV-ID is compatible with the presence of paper in the reaction tube and suggest that self-collection of saliva by patients directly on absorbent paper could provide a simple and cost-effective strategy to collect and test thousands of saliva samples for multiple pathogens (**Fig. 4F**).

## DISCUSSION

Testing strategies are vital to an effective public health response to the COVID-19 pandemic, particularly with the spread of the disease by asymptomatic individuals. An ongoing challenge to COVID-19 testing is the need for massive testing strategies for population-level surveillance that are needed for efficient contact tracing and isolation. Most FDA-approved clinical SARS-CoV-2 diagnostic tests are based on time-consuming and expensive protocols that include RNA purifications and RT-PCR^32^ and must be performed by trained personnel in well-equipped laboratories. Point-of-care antigen tests provide a much faster turnaround time and require little manipulation, but there remains limited data on their specificity in real-world applications^40^. Because of reagent limitations and diagnostic testing bottlenecks, prioritization of COVID diagnostic testing continues to be for symptomatic individuals and individuals who are particularly vulnerable for infection after exposure^41^. Private organizations, including colleges and universities, have circumvented some of these challenges by contracting with private laboratories to establish asymptomatic surveillance testing protocols; this is a costly option for population-level surveilling of asymptomatic SARS-CoV-2 infections.

Several effective COVID-19 vaccines have been developed and there is a concerted ongoing global vaccination effort, providing a concrete means to end the pandemic. Despite this progress there are several potential risks that require vigilance: possible COVID-19 transmission in vaccinated individuals, emergence of vaccine-resistant viral variants, and public skepticism of vaccines or faltering compliance with social distancing guidelines^42^. For these reasons ongoing testing and surveillance efforts will remain important for the foreseeable future, both to monitor the progress of vaccination in reducing symptomatic cases and to detect emerging variants.

In order to scale testing to an effective volume and frequency, surveillance tests must demonstrate the following qualities: 1) <u>sensitivity</u>, to identify both asymptomatic and symptomatic carriers; 2) <u>simplicity</u> in methodology, to be performed in a number of traditional diagnostic laboratories, without specialized equipment; 3) <u>low cost</u> and easily accessible reagents; 4) <u>ease of collection</u> method; 5) <u>rapid</u> turnaround time to allow for isolation and contract tracing; and 6) ability to <u>co-detect</u> multiple respiratory viruses, given the overlap in patient symptoms. To this end, we have developed COV-ID, an RT-LAMP-based parallel sequencing SARS-CoV-2 detection method that can provide results from tens of thousands of samples per day at relatively low cost to simultaneously detect multiple respiratory viruses.

COV-ID features several key innovations that make it well-suited to high-throughput testing. First, COV-ID uses a two-dimensional barcoding strategy^8^, where the same 96 barcodes are used in each RT-LAMP plate, making it possible to pre-aliquot barcodes in 96-well plates ahead of time and store them at −20ºC, simplifying execution of the assay and shortening turnaround times. Second, since RT-LAMP does not require thermal cycling, tens of thousands of samples can be run simultaneously in a standard benchtop-sized incubator or hybridization oven held at 65ºC. Third, individual samples are pooled immediately following RT-LAMP; therefore, a single thermocycler has the potential to process up to 96 or 384 RT-LAMP plates, generating 9,216 or 36,864 individually barcoded samples, respectively (**Fig. 1A, 4F, 4G**). Only 96 unique FIP barcodes are required for this scaling; here, we show that 28 out of 32 LAMP barcodes tested were functional for both *N2* and *STATH*. This proof-of-principle experiment demonstrates the feasibility of generating the library of barcodes required to apply COV-ID to a large population. An additional advantage of sequencing-based approaches, such as COV-ID is that with carefully designed primers it would be possible to recover information about viral variants directly from the sequencing reads^43^. Finally, COV-ID can generate ready-to-sequence libraries directly from saliva absorbed onto filter paper, which would allow for major streamlining of the often-challenging logistical process of sample collection (**Fig. 4**). Thus, COV-ID libraries for thousands and tens of thousands of samples can be generated with relatively minimum effort in biological laboratories with basic equipment and easily accessible reagents.

With the average throughput of an Illumina NextSeq 500/550, a relatively affordable next-generation sequencer up to 9,216 (96 RT-LAMPs x 96 pools) can be sequenced at a depth of ∼48,000 reads per sample, and up to 36,864 (96 RT-LAMPs x 384 pools) can be sequenced at a depth of ∼12,000 reads, which, we showed, is more than sufficient to obtain information about multiple viral and control amplicons. Considering that reagents for one NextSeq run cost ∼1,500 U.S. dollars, the theoretical sequencing cost per sample could be as low as $0.04 (**Fig. 4G**). While sequencing instruments are relatively specialized and not ubiquitous, amplified COV-ID DNA libraries could be shipped to remote facilities for sequencing in a cost-effective manner as previously proposed by the inventors of LAMP-seq^14^. Finally, because of the limited sequence space against which reads must be aligned, computational analysis of the resulting data can be performed in a matter of minutes with optimized pipelines, providing results shortly after the sequencing run has completed.

COV-ID has sensitivity of 5–10 virions of SARS-CoV-2 per μL in contrived saliva samples (**Fig. 2D**) and at least 300 virions / μL in saliva collected from patients in a clinical setting (**Fig. 2E**). However, this was sufficient to properly classify 100% of the clinical samples analyzed, given that all positive samples had an estimated viral load > 300 virions / µL. Importantly, this was also the apparent limit of sensitivity of paper-based COV-ID (**Fig. 4E**), suggesting that even in these settings COV-ID would be capable of accurately classifying the majority of patient-derived samples.

In conclusion, COV-ID is a flexible platform that can be executed at varying levels of scale with additional flexibility in sample input, making it an attractive platform for surveillance testing. Population-level monitoring of SARS-CoV-2 infections will be critical while vaccines are being distributed to the global population, and continued surveillance will likely remain an effective strategy to protect immune-compromised and unvaccinated members in society and within entities and organizations where regular monitoring is critical to social isolation strategies. To that end, effective, low-cost, multiplexed, and readily-implementable strategies for surveillance testing, such as COV-ID, are important to mitigate the effects of the current and future pandemics.

## METHODS

### RT-LAMP primer design

Primers against *ACTB* were designed using PrimerExplorerV5 (https://primerexplorer.jp/e/) using default parameters and including loop primers (**Table S1**).

For COV-ID, priming sequences for PCR were inserted in FIP and BIP primers between the target homology regions (F1c and F2, and B1c and B2, respectively, see **Fig. S1**). After testing, we determined that 12 nts and 11 nts were most effective for the P5 and P7 binding regions, respectively, being the shortest insertion that allowed reliable PCR amplification from LAMP products without impacting LAMP efficiency. In addition a 5 nt barcode sequence was inserted at the immediate 3’ end of the P5-binding region of the FIP primer.

### LAMP barcode design

Starting from the total possible 1,024 unique 5-nt barcodes, we removed those that matched any sequence within the RT-LAMP primers used in this study (**Table S1**) in either sense or anti-sense orientation. From the remaining pool, we selected 32 barcodes with hamming distance of at least 2 between all candidates. We tested FIPs incorporating candidate barcodes for *ACTB, STATH, N2*, and IAV primer sets on saliva RT-LAMP with 1,000 copies target amplicon. Primers that failed to show LAMP signal by real time fluorescence monitoring or generate expected PCR product were discarded. Final usable barcodes are provided in (**Table S2**).

### Saliva preparation

We prepared 100x TCEP/EDTA buffer (250 mM TCEP, 100 mM EDTA, 1.15 N NaOH) ^29^. TCEP/EDTA buffer was added to human saliva at 1:100 volume, then samples were capped, vortexed to mix and heated in a thermocycler (95ºC 5 min, 4ºC hold) until ready to use for RT-LAMP. When indicated, heat-inactivated SARS-CoV-2 (BEI Resources Cat. NR-52286) or H1N1 genomic RNA (Twist Biosciences Cat. 103001) was added to inactivated saliva prior to RT-LAMP.

### N2 spike-in synthesis

To prepare the *in vitro* transcription template for SARS-CoV-2 *N2* spike-in RNA, we performed RT-PCR using Power SYBR RNA-to-Ct kit (Thermo Cat. 4389986) of heat inactivated SARS-CoV-2 (BEI Resources Cat. NR-52286) using the following primers: N2-B3 and N2-spike-T7 S. PCR product was purified and used as a template for in vitro transcription using HiScribe T7 transcription kit (NEB Cat. E2040S). RNA was purified with Trizol (Thermo Cat. 15596026), quantified via A_260_, then aliquoted in BTE buffer (10 mM bis-tris pH 6.7, 1 mM EDTA) and stored at −80ºC. Primers used and final spike-in sequence are provided in **Table S1**.

### RT-LAMP

All RT-LAMP reactions were set up in clean laminar flow hoods and all steps before and after LAMP were carried out in separate lab spaces to avoid contamination. RT-LAMP reactions were set up on ice as follow: for each amplicon 5 or 6 LAMP primers were combined into 10x working stock at established concentrations: 16 μM FIP, 16 μM BIP, 4 μM LF, 4 μM LB, 2 μM F3, 2 μM B3. For multiplexed COV-ID reactions 10x working primer mixes for each amplicon were either added proportionally so that the total primer content remained constant, or mixed so that BIP and FIP primers were scaled down depending on amplicon number while remaining primers (LF and/or LB, F3, B3) were kept at same concentration as in single reactions.

Each 10 μL RT-LAMP reaction mix consisted of 1x Warmstart LAMP 2x Master Mix (NEB Cat. E1700S), 0.7 μM dUTP (Promega Cat. U1191), 1 μM SYTO-9 (Thermo Cat. S34854), 0.1 μL Thermolabile UDG (Enzymatics Cat. G5020L), 1 μL of saliva template and optionally 20 fg of N2 Spike RNA. Reactions were prepared in qPCR plates or 8-well strip tubes, sealed, vortexed and centrifuged briefly, then incubated in either a QuantStudio Flex 7 or StepOnePlus instrument (Thermo) for 65ºC 1 hr. Real-time fluorescence measurements were recorded every 30 sec to monitor reaction progress but were not used for data analysis. Following LAMP the reactions were heated at 95ºC 5 min to inactivate LAMP enzymes.

### Library construction by PCR amplification

All post-LAMP steps were carried out on a clean bench separate from LAMP reagents and workspace. For individual LAMP samples, LAMP amplicons were diluted either 1:100 or 1:1,000 in water. For pooling of individually barcoded LAMP reactions, equal amounts of all LAMP reactions were combined and then either diluted 1:1000 or purified via SPRIselect beads (Beckman Coulter Cat. B23317) using a bead-to-reaction ratio of 0.1x. Purified material was diluted to final 100-fold dilution relative to LAMP.

1 μL of diluted LAMP material was used as a template for PCR using OneTaq DNA polymerase (NEB Cat. M0480L) with 100 nM each of custom dual-indexed Illumina P5 and P7 primers in either 10 or 25 μL reaction (**Table S1**). PCR reactions were incubated as follows: (25 cycles of stage 1 [94ºC x 15 sec, 45ºC x 15 sec, 68ºC x 10 sec], 10 cycles of Stage 2 [94ºC x 15 sec, 68ºC x 10 sec], 68ºC x 1 min, 4ºC x ∞). Note, for initial pilot COV-ID and clinical sample experiments (**Fig. 2D–E, Fig. S2C**) PCR incubation was performed as above with modification: [Stage 1 × 10 cycles, Stage 2 × 25 cycles].

PCR products were resolved on 2% agarose gel to confirm library size, then all were pooled and purified via MinElute PCR purification kit (Qiagen Cat. 28004) and quantified using either Qubit dsDNA High Sensitivity kit (Thermo Cat. Q32851) or Kapa Library Quantification Kit for Illumina (Kapa Cat. 07960140001).

### Human samples

Clinical saliva samples used for **Fig. 2E** were obtained and characterized as part of a separate study at the University of Pennsylvania^44^ and collected under Institutional Review Board (IRB)-approved protocols (IRB protocol #842613 and #813913). Briefly, salivary samples were collected from possible SARS-CoV-2 positive patients at one of three locations: (1) Penn Presbyterian Medical Center Emergency Department, (2) Hospital of the University of Pennsylvania Emergency Department, and (3) Penn Medicine COVID-19 ambulatory testing center. Inclusion criteria including any adult (age > 17 years) who underwent SARS-CoV-2 testing via standard nasopharyngeal swab at the same visit. Patients with known COVID-19 disease who previously tested positive previously were excluded. After verbal consent was obtained by a trained research coordinator, patients were instructed to self-collect saliva into a sterile specimen container which was then placed on ice until further processing for analysis.

The saliva used in the remaining experiments was donated by one of the authors. Because it was only used for protocol optimization the Penn IRB has determined that it did not constitute human subjects research and therefore approval was not required.

### Paper COV-ID

Squares of Whatman no. 1 filter paper (2 mm x 2 mm) were cut using a scalpel on a clean surface under a laminar flow hood and stored at room temperature until used. Using ethanol-sterilized fine-nosed tweezers a single square was dipped twice into unprocessed, freshly collected saliva with or without added SARS-CoV-2 (BEI Resources Cat. NR-52286) until saliva was saturated on paper by eye. Paper was then transferred to well of 96-well plate containing 10 ul of 1x TCEP/EDTA buffer (2.5 mM TCEP, 1 mM EDTA, 1.15 NaOH). Plate was placed on heat block inside laminar flow hood or inside open thermocycler and incubated at 95ºC x 10 min.

10 ul RT-LAMP mixture was prepared as described above in the absence of the N2 Spike RNA. 10 ul of RT-LAMP reaction mixture was added to each paper strip, then plate was sealed and incubated 65ºC x 1 hr, 95ºC x 5 min in QuantStudio Flex 7 (Thermo). 1 ul of each reaction was either diluted 1:100 or purified via SPRIselect beads and PCR amplified as described above.

### Sequencing

Libraries were sequenced on one of the following Illumina instruments: MiSeq, NextSeq 500, NextSeq 550, NovaSeq 6000 and sequenced using single end programs with a minimum of 40 cycles on Read 1 and 8 cycles for index 1 (on P7) and index 2 (on P5).

### Sequence Analysis

Reads were filtered for optical quality using FASTX-toolkit utility fastq_quality_filter (http://hannonlab.cshl.edu/fastx_toolkit/), then cutadapt^45^ was used to remove adapters and demultiplex LAMP barcodes. Reads were aligned to a custom index containing SARS-CoV-2 genome (NC_045512.2), Influenza H1N1 coding sequences (NC_026431.1, NC_026432.1, NC_026433.1, NC_026434.1, NC_026435.1, NC_026436.1, NC_026437.1, NC_026438.1), STATH coding sequence (NM_003154.3), and custom N2 spike sequence (**Table S3**), target sequences using bowtie2^46^ with options --no-unal and --end-to-end. Alignments with greater than 1 mismatch were removed and the number of reads mapping to each target for all barcodes were extracted and output in a matrix. Barcodes with fewer than 25 total mapped reads were discarded.

## Data Availability

Next generation sequencing data generated for this study are available at the NCBI GEO with accession GSE172118.

https://www.ncbi.nlm.nih.gov/geo/query/acc.cgi?acc=GSE172118

## ACKNOWLEDGMENTS

The authors thank E. Shields for careful proofreading of analysis scripts; B. Morris and R. Collman for the collection and distribution of clinical saliva samples; F. Bushman, S. Sherril-Mix, and Abigail Glascock for sharing RT-qPCR data on the clinical samples; the UPenn rapid assay task force for project feedback; and the gLAMP weekly forum for advice and guidance.

## DISCLOSURE

R.W-T., C.A.T. and R.B. are inventors on a patent application filed by the University of Pennsylvania related to this work.

## SUPPLEMENTARY FIGURE LEGENDS

**Figure S1.**
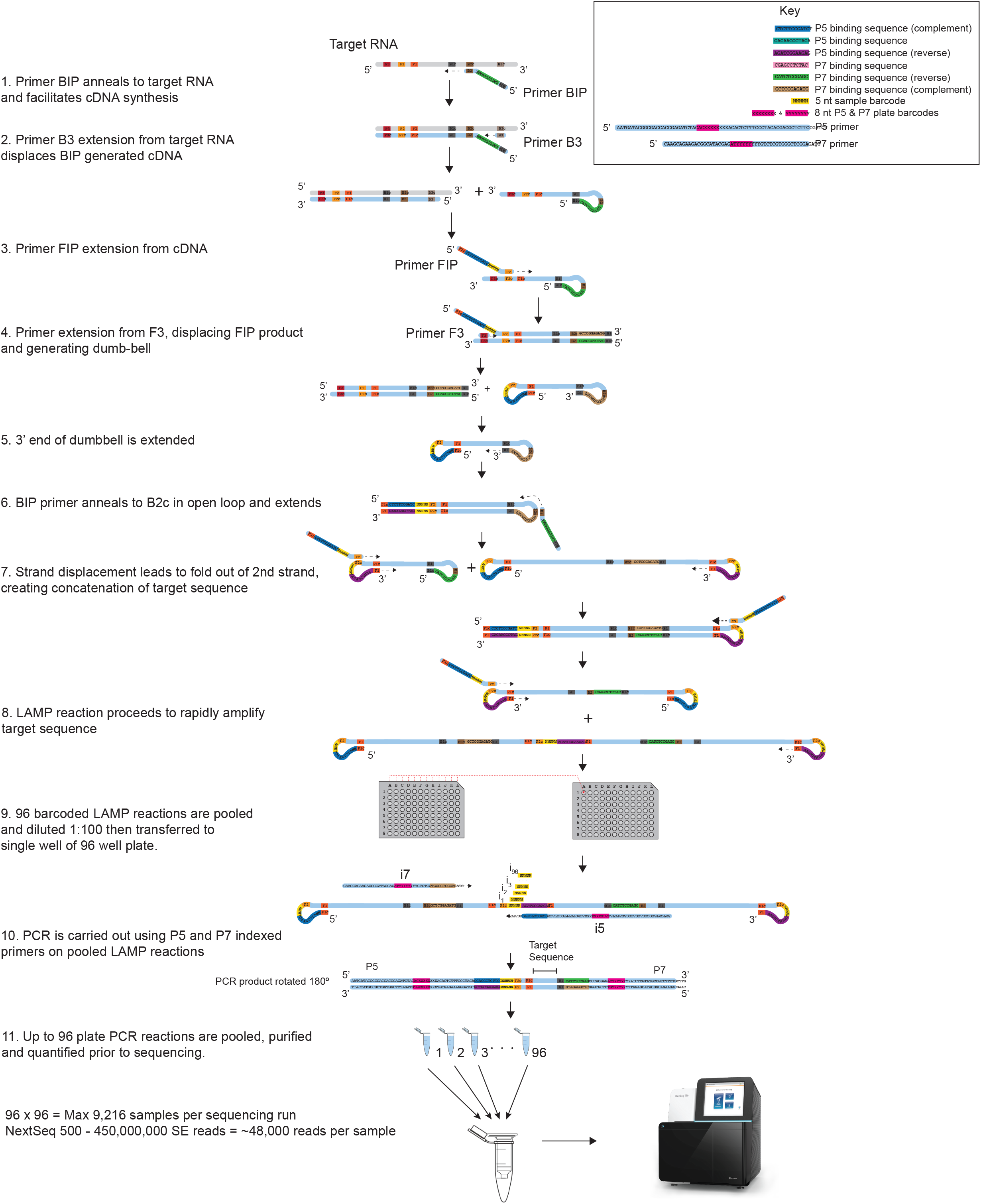
Detailed COV-ID mechanism. The steps of the COV-ID protocol are depicted, showing RT-LAMP mechanism and the final barcoded amplicon that is sequenced. For clarity only, selected steps of RT-LAMP reaction are shown and loop primer intermediates are not depicted. For full LAMP mechanism see (Nagamine et al., 2002).

**Figure S2.**
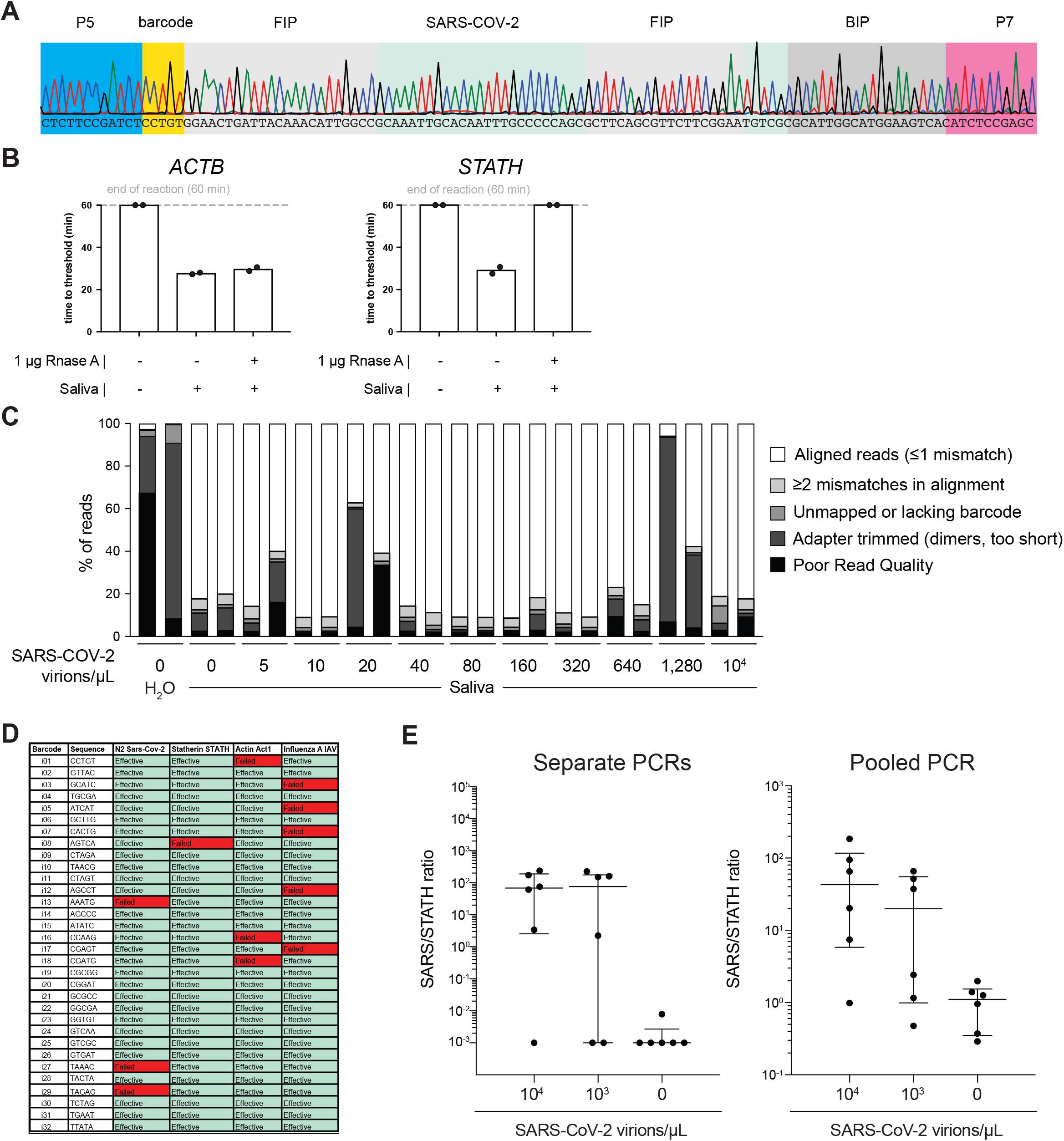
Optimization of COV-ID in human saliva. (A) Saliva COV-ID sequence validation. Single saliva COV-ID reaction using N2 primers was sequenced by the Sanger method. (B) Validation of control human amplicons for RT-LAMP on saliva. RT-LAMP of TCEP/EDTA inactivated saliva was performed with conventional RT-LAMP primer sets for ACTB and STATH in the presence or absence of RNase A. (C) Characterization of COV-ID sequencing libraries. Breakdown of reads for sequence data presented in Fig. 2D. Samples without added template consist of predominantly adapter dimers. (D) Validation of COV-ID LAMP barcodes. 32 potential barcodes were tested for LAMP primer sets indicated, incompatible barcodes are marked in red. (E) Validation of pooled PCR. COV-ID was performed on saliva samples using unique LAMP barcodes. The RT-LAMP reactions were then amplified either by individual PCR or by first pooling and then performing a single PCR on the pool.

**Figure S3.**
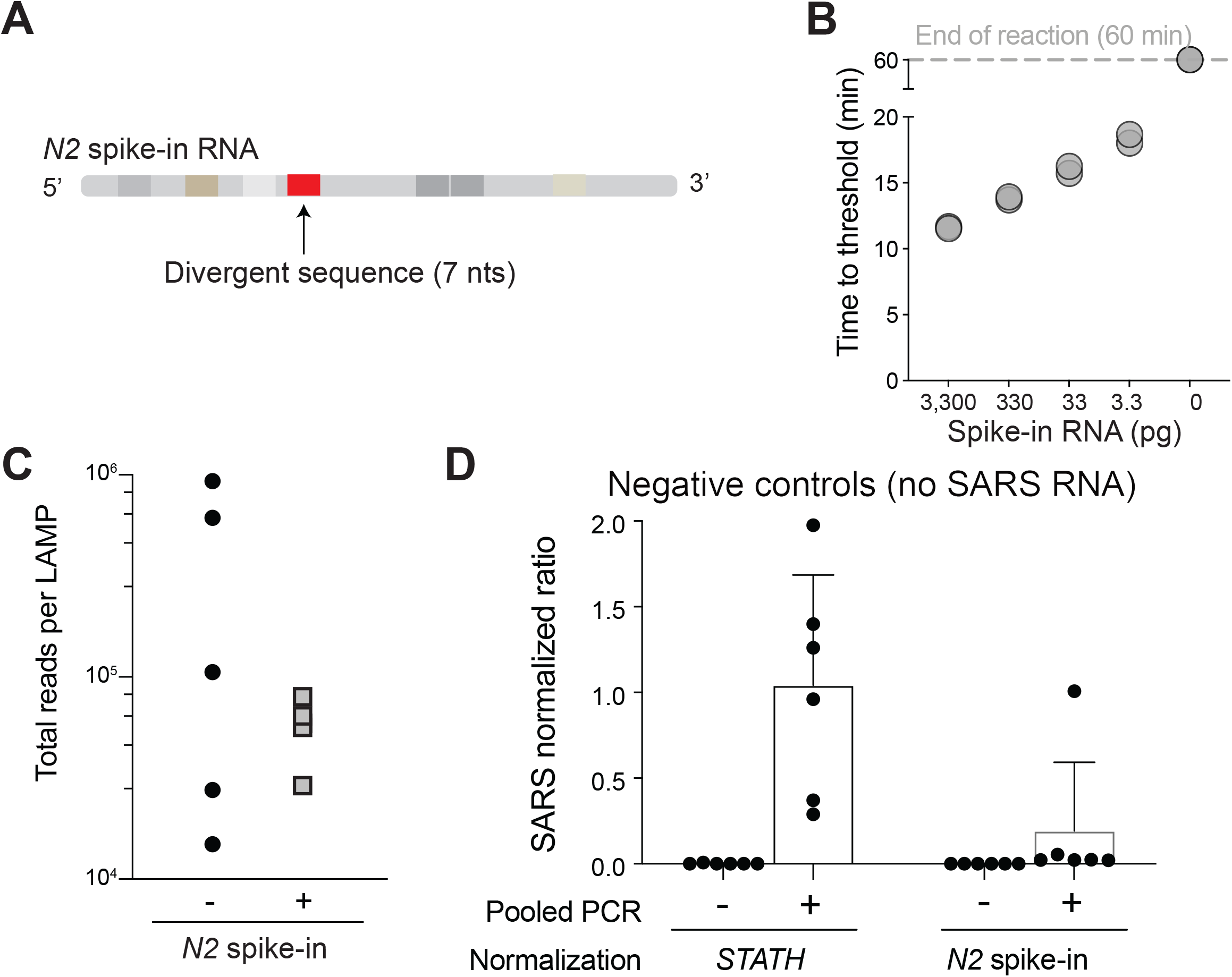
Spike-in strategy for COV-ID’. (A) Synthetic *N2* Spike RNA. SARS-CoV-2 *N2* RNA fragment was synthesized including 7 nt divergent sequence inside the forward loop primer-binding site, maintaining all other LAMP primer binding sites and identical GC content. (B) RT-LAMP using COV-ID *N2* primers was carried out on indicated amounts of spike-in RNA, showing rapid amplification down to picogram quantities of added template. (C) Total number of reads per barcode in COV-ID pool obtained by including (+) or omitting (-) the *N2* spike-in. (D) Spurious COV-ID signal for the *N2* amplicon in negative control samples after normalization either to the *STATH* control in absence of spike-in (left) or to the *N2* spike-in control.

## Notes

### Funding Statement

This work was internally funded in part by the Rapid Assay Task Force initiative at the University of Pennsylvania.

### Author Declarations

Clinical saliva samples used for Fig. 2E were obtained and characterized as part of a study at the University of Pennsylvania and collected under Institutional Review Board (IRB)-approved protocols (IRB protocol #842613 and #813913). The saliva used in the remaining experiments was donated by one of the authors. Because it was only used for protocol optimization the Penn IRB has determined that it did not constitute human subjects research and therefore approval was not required.

## REFERENCES

1. Haug, N. et al.. Ranking the effectiveness of worldwide COVID-19 government interventions. Nat Hum Behav 4, 1303–1312 (2020).

2. Tian, H. et al.. An investigation of transmission control measures during the first 50 days of the COVID-19 epidemic in China. Science 368, 638–642 (2020).

3. Taipale, J., Romer, P. & Linnarsson, S. Population-scale testing can suppress the spread of COVID-19. medRxiv, 2020.2004.2027.20078329 (2020).

4. Endo, A., Centre for the Mathematical Modelling of Infectious Diseases, C.-W.G., Abbott, S., Kucharski, A.J. & Funk, S. Estimating the overdispersion in COVID-19 transmission using outbreak sizes outside China. Wellcome Open Res 5, 67 (2020).

5. Holt, E. Slovakia to test all adults for SARS-CoV-2. Lancet 396, 1386–1387 (2020).

6. Larremore, D.B. et al.. Test sensitivity is secondary to frequency and turnaround time for COVID-19 screening. Sci Adv (2020).

7. Bloom, J.S. et al.. Swab-Seq: A high-throughput platform for massively scaled up SARS-CoV-2 testing. medRxiv, 2020.2008.2004.20167874 (2020).

8. Yelagandula, R. et al.. SARSeq, a robust and highly multiplexed NGS assay for parallel detection of SARS-CoV2 and other respiratory infections. medRxiv, 2020.2010.2028.20217778 (2020).

9. Aynaud, M.-M. et al.. A Multiplexed, Next Generation Sequencing Platform for High-Throughput Detection of SARS-CoV-2. medRxiv, 2020.2010.2015.20212712 (2020).

10. Wu, Q. et al.. INSIGHT: A population-scale COVID-19 testing strategy combining point-of-care diagnosis with centralized high-throughput sequencing. Sci Adv 7 (2021).

11. Chappleboim, A. et al.. ApharSeq: An Extraction-free Early-Pooling Protocol for Massively Multiplexed SARS-CoV-2 Detection. medRxiv, 2020.2008.2008.20170746 (2020).

12. James, P. et al.. LamPORE: rapid, accurate and highly scalable molecular screening for SARS-CoV-2 infection, based on nanopore sequencing. medRxiv, 2020.2008.2007.20161737 (2020).

13. Dao Thi, V.L. et al. A colorimetric RT-LAMP assay and LAMP-sequencing for detecting SARS-CoV-2 RNA in clinical samples. Sci Transl Med 12 (2020).

14. Schmid-Burgk, J.L. et al.. LAMP-Seq: Population-Scale COVID-19 Diagnostics Using a Compressed Barcode Space. bioRxiv, 2020.2004.2006.025635 (2020).

15. Li, S. et al.. Simultaneous detection and differentiation of dengue virus serotypes 1-4, Japanese encephalitis virus, and West Nile virus by a combined reverse-transcription loop-mediated isothermal amplification assay. Virol J 8, 360 (2011).

16. Shirato, K. et al.. Detection of Middle East respiratory syndrome coronavirus using reverse transcription loop-mediated isothermal amplification (RT-LAMP). Virol J 11, 139 (2014).

17. Calvert, A.E., Biggerstaff, B.J., Tanner, N.A., Lauterbach, M. & Lanciotti, R.S. Rapid colorimetric detection of Zika virus from serum and urine specimens by reverse transcription loop-mediated isothermal amplification (RT-LAMP). PLoS One 12, e0185340 (2017).

18. Enomoto, Y. et al.. Rapid diagnosis of herpes simplex virus infection by a loop-mediated isothermal amplification method. J Clin Microbiol 43, 951–955 (2005).

19. Augustine, R. et al.. Loop-Mediated Isothermal Amplification (LAMP): A Rapid, Sensitive, Specific, and Cost-Effective Point-of-Care Test for Coronaviruses in the Context of COVID-19 Pandemic. Biology (Basel) 9 (2020).

20. United States Food and Drug Administration. Color Genomics SARS-CoV-2 RT-LAMP Diagnostic Assay - EUA Summary. https://www.fda.gov/media/138249/download. Published: November 2, 2020, Accessed: December 22, 2020.

21. Nagamine, K., Hase, T. & Notomi, T. Accelerated reaction by loop-mediated isothermal amplification using loop primers. Mol Cell Probes 16, 223–229 (2002).

22. Notomi, T. et al.. Loop-mediated isothermal amplification of DNA. Nucleic Acids Res 28, E63 (2000).

23. Yamagishi, J. et al.. Serotyping dengue virus with isothermal amplification and a portable sequencer. Sci Rep 7, 3510 (2017).

24. Butler, D.J. et al.. Shotgun Transcriptome and Isothermal Profiling of SARS-CoV-2 Infection Reveals Unique Host Responses, Viral Diversification, and Drug Interactions. bioRxiv, 2020.2004.2020.048066 (2020).

25. Savela, E.S. et al.. SARS-CoV-2 is detectable using sensitive RNA saliva testing days before viral load reaches detection range of low-sensitivity nasal swab tests. medRxiv, 2021.2004.2002.21254771 (2021).

26. Ranoa, D.R.E. et al.. Saliva-Based Molecular Testing for SARS-CoV-2 that Bypasses RNA Extraction. bioRxiv, 2020.2006.2018.159434 (2020).

27. Myhrvold, C. et al.. Field-deployable viral diagnostics using CRISPR-Cas13. Science 360, 444–448 (2018).

28. Lalli, M.A. et al.. Rapid and extraction-free detection of SARS-CoV-2 from saliva with colorimetric LAMP. medRxiv, 2020.2005.2007.20093542 (2020).

29. Rabe, B.A. & Cepko, C. SARS-CoV-2 detection using isothermal amplification and a rapid, inexpensive protocol for sample inactivation and purification. Proc Natl Acad Sci U S A 117, 24450–24458 (2020).

30. Babiker, A., Myers, C.W., Hill, C.E. & Guarner, J. SARS-CoV-2 Testing. Am J Clin Pathol 153, 706–708 (2020).

31. Satoh, T. et al.. Development of mRNA-based body fluid identification using reverse transcription loop-mediated isothermal amplification. Anal Bioanal Chem 410, 4371–4378 (2018).

32. MacKay, M.J. et al.. The COVID-19 XPRIZE and the need for scalable, fast, and widespread testing. Nat Biotechnol 38, 1021–1024 (2020).

33. Torres, C. et al.. LAVA: an open-source approach to designing LAMP (loop-mediated isothermal amplification) DNA signatures. BMC Bioinformatics 12, 240 (2011).

34. Zhang, Y. & Tanner, N.A. Development of Multiplexed RT-LAMP for Detection of SARS-CoV-2 and Influenza Viral RNA. medRxiv, 2020.2010.2026.20219972 (2020).

35. Takayama, I. et al.. Development of real-time fluorescent reverse transcription loop-mediated isothermal amplification assay with quenching primer for influenza virus and respiratory syncytial virus. J Virol Methods 267, 53–58 (2019).

36. Glushakova, L.G. et al.. Detection of chikungunya viral RNA in mosquito bodies on cationic (Q) paper based on innovations in synthetic biology. J Virol Methods 246, 104–111 (2017).

37. Kellner, M.J. et al.. A rapid, highly sensitive and open-access SARS-CoV-2 detection assay for laboratory and home testing. bioRxiv, 2020.2006.2023.166397 (2020).

38. Yaren, O. et al.. Ultra-rapid detection of SARS-CoV-2 in public workspace environments. medRxiv, 2020.2009.2029.20204131 (2020).

39. Winnett, A. et al.. SARS-CoV-2 Viral Load in Saliva Rises Gradually and to Moderate Levels in Some Humans. medRxiv (2020).

40. Pettengill, M.A. & McAdam, A.J. Can We Test Our Way Out of the COVID-19 Pandemic? J Clin Microbiol 58 (2020).

41. Schuetz, A.N. et al.. When Should Asymptomatic Persons Be Tested for COVID-19? J Clin Microbiol 59 (2020).

42. Aschwanden, C. Five reasons why COVID herd immunity is probably impossible. Nature 591, 520–522 (2021).

43. Everett, J. et al.. SARS-CoV-2 Genomic Variation in Space and Time in Hospitalized Patients in Philadelphia. mBio 12 (2021).

44. Sherrill-Mix, S. et al.. LAMP-BEAC: Detection of SARS-CoV-2 RNA Using RT-LAMP and Molecular Beacons. medRxiv, 2020.2008.2013.20173757 (2020).

45. Martin, M. Cutadapt removes adapter sequences from high-throughput sequencing reads. 2011 17, 3 (2011).

46. Langmead, B. & Salzberg, S.L. Fast gapped-read alignment with Bowtie 2. Nat Methods 9, 357–359 (2012).

